# Epigenomic Aberrations of Histone Methylation in Prefrontal Cortex of Humans with Mild Cognitive Impairment and Alzheimer’s Disease

**DOI:** 10.1101/2025.07.17.25331659

**Authors:** Prachetas Jai Patel, Zhen Yan

**Affiliations:** Department of Physiology and Biophysics, State University of New York (SUNY) at Buffalo, School of Medicine and Biomedical Sciences, 955 Main Street, Buffalo, NY, 14203; VA Western New York Healthcare System, 3495 Bailey Avenue, Research Building 20, Buffalo, NY 14215

## Abstract

Epigenetic mechanisms, particularly histone modifications at gene promoters, are crucial for controlling gene transcription. During the progression of neurodegenerative disorders, epigenomic aberrations may contribute to gene dysregulation, leading to manifestation of symptoms. To test this, we employed a multifaceted approach to investigate how the two key histone methylation marks, H3K4me3 (linked to gene activation) and H3K27me3 (linked to gene suppression), are altered in postmortem prefrontal cortex of humans with Mild Cognitive Impairment (MCI) or Alzheimer’s Disease (AD). Compared to controls, MCI and AD exhibited pronounced losses of permissive H3K4me3 peaks at promoters of genes enriched in synaptic plasticity and neurotransmission, and significant gains of H3K4me3 peaks at promoters of genes enriched in transcriptional regulation. AD displayed more substantial H3K4me3 losses on synaptic genes than MCI. Conversely, significant gains of repressive H3K27me3 peaks were observed at synaptic gene promoters in both disease groups, with MCI exhibiting more pronounced H3K27me3 gains on synaptic genes than AD. Weighted Gene Correlation Network Analysis (WGCNA) revealed multiple modules characterizing distinct patterns of gains and losses of H3K4me3 and H3K27me3 during the transition from MCI to AD. Integrative analysis of epigenomic and transcriptomic data indicated that these histone mark alterations were well correlated with the downregulation of synaptic genes and upregulation of transcriptional regulators in AD. This comprehensive profiling uncovers a stage-dependent reorganization of histone modifications at critical gene loci, implicating these events in the molecular cascade of AD pathogenesis. Targeting dysregulated chromatin states may offer novel therapeutic avenues for early intervention of AD.

## Introduction

Alzheimer’s disease (AD) is primarily characterized by progressive cognitive decline. Despite extensive research, the mechanisms driving the disease’s onset and progression remain elusive, limiting therapeutic interventions primarily to symptom management targeting hallmark pathological features, such as amyloid-beta plaques, tau neurofibrillary tangles, and inflammatory responses (Karran & De Strooper 2022, Wilson et al 2023). Consequently, there is a compelling need to identify underlying molecular events that causally link to AD pathology.

Emerging evidence strongly implicates epigenetic aberrations, particularly histone modification changes, as significant contributors to the aging process and neurodegenerative disorders (Berson et al 2018, Lardenoije et al 2015, López-Otín et al 2023, Santana et al 2023, Wilson et al 2023). Histone modifications, such as histone acetylation and methylation, play a critical role in gene transcription through regulating chromatin structure and accessibility, profoundly impacting cellular function (Kouzarides 2007, Santana et al 2023, Smith & Shilatifard 2010). While the alteration of histone acetylation in AD has been well documented (Gräff et al 2012, Klein et al 2019, Marzi et al 2018, Nativio et al 2018, Nativio et al 2020), changes in histone methylation in AD is much less known. The histone H3 trimethylation at lysine 4 (H3K4me3) and at lysine 27 (H3K27me3) is strongly associated with transcriptional activation and repression, respectively (Cai et al 2021, Wang et al 2023). However, it is unclear whether these key epigenetic marks are dysregulated in neurodegenerative conditions.

The current study seeks to reveal the alterations in H3K4me3 and H3K27me3 at gene promoters in the prefrontal cortex of humans with Mild Cognitive Impairment (MCI) or AD. MCI is often considered a precursor or a transitional state into AD (Morris et al 2001). Hence, understanding the convergent epigenomic mechanisms in the two disease states could provide significant mechanistic insights.

We implemented a novel integrative strategy combining genome-wide ChIP-seq and Weighted Gene Co-expression Network Analysis (WGCNA) to elucidate co-binding patterns of histone modifications (H3K4me3 and H3K27me3) across promoters during the transition from MCI to AD. This method uniquely reveals how coordinated changes in histone marks contribute to disease progression, a previously unexplored angle in neuroepigenomics.

Furthermore, we integrated epigenomic data with transcriptomic profiles using network-based approaches, thereby elucidating critical molecular pathways that are altered in MCI and AD. Our findings presented herein underscore the significance of epigenomic disruptions in the progression of AD from MCI and highlight histone modifications as promising targets for therapeutic strategies aimed at early intervention and potentially AD prevention.

## Results

### H3K4me3 Occupancies Are Lost on Synaptic Genes and Gained on Transcription Regulators in Both MCI and AD

H3K4me3 ChIP-seq data were examined in 11 MCI, 17 AD, and 11 age-matched control samples with no cognitive impairment. As shown in the PCA plots (**Fig. 1A**), the diseased groups were largely segregated from controls in the sample distribution of H3K4me3 peaks. Differential binding analysis identified many significantly changed (Fold ≥ |1.1|, p-value ≤ 0.05) genomic sites with H3K4me3 occupancy (MCI: 4368; AD: 3319). Filtering and limiting to the promoter region (+/− 3kb of TSS) of protein-coding genes, MCI had significant H3K4me3 gains on 436 sites (407 genes) and losses on 720 sites (697 genes), while AD had significant H3K4me3 gains on 564 sites (534 genes) and losses on 702 sites (675 genes) (**Fig. 1B, Sup. Table 1**).

**Figure 1.**
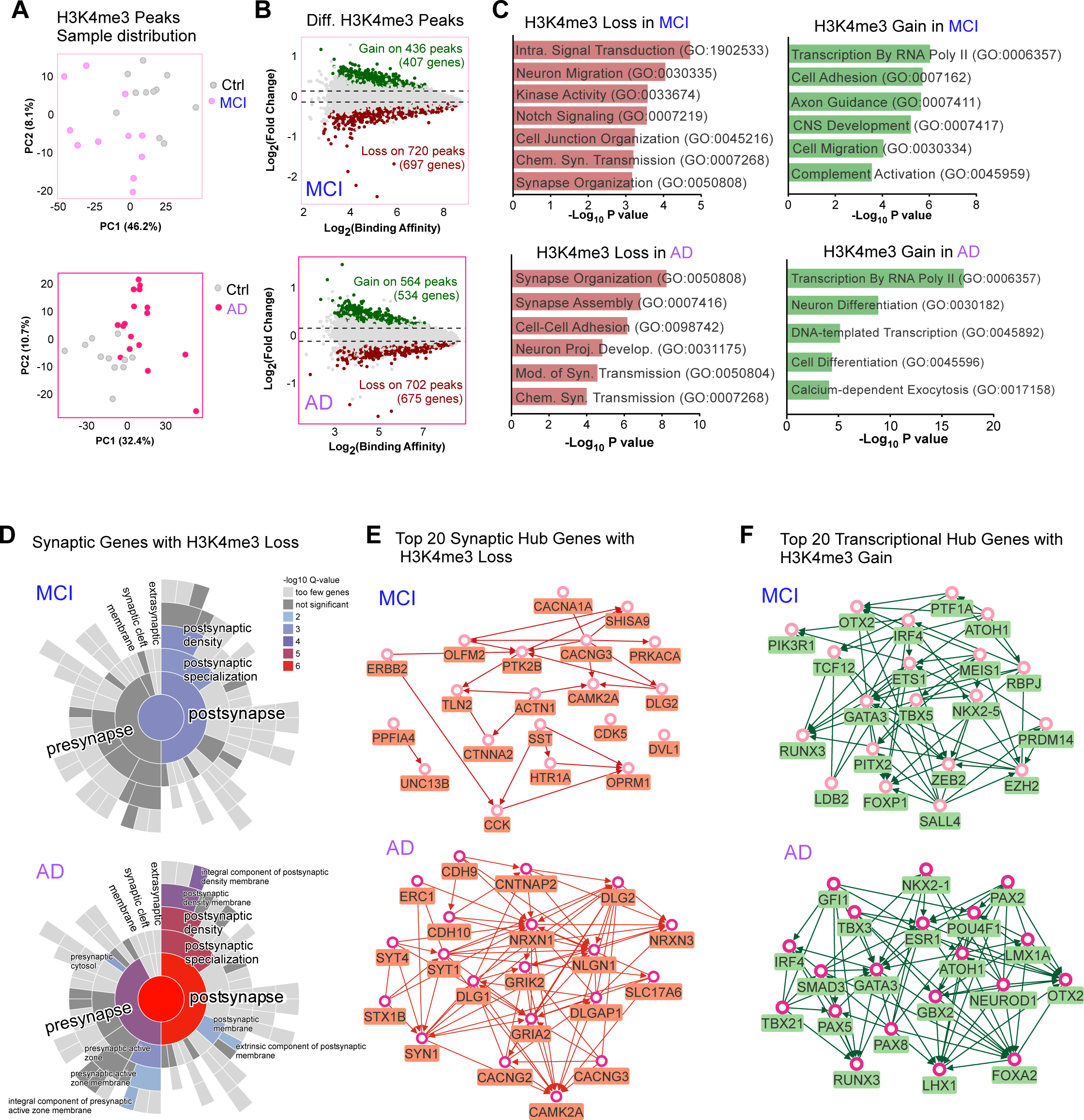
H3K4me3 Alterations in MCI and AD. **A,** PCA plots depicting H3K4me3 peaks’ sample distribution in control (Ctrl) vs. MCI (top) or Ctrl vs. AD (bottom). **B,** MA plots depicting differential H3K4me3 binding scores in MCI (top) and AD (below), compared to Ctrl. Significant gains (green) and losses (red) are also labeled. P-values were calculated using a two-sided Wilcoxon ‘Mann-Whitney’ test, with a threshold of ≤ 0.05 **C,** Gene Ontology (GO) of protein-coding genes with significant H3K4me3 losses (left) or gains (right) at their promoters in MCI (top) and AD (bottom). P-values were calculated using Fisher’s Exact test. **D,** SYNGO cellular component analysis of synaptic genes with significant H3K4me3 losses in MCI (top) and AD (bottom). P-values were calculated using Fisher’s Exact test. **E,** PPI of top 20 synaptic hub genes with H3K4me3 losses in MCI (top) and AD (bottom). **F,** PPI of top 20 transcriptional hub genes with H3K4me3 gains in MCI (top) and AD (bottom).

Using Gene Set Enrichment analysis (GSEA), we examined Gene Ontology (GO) pathways of these genes with significant changes in H3K4me3 binding at promoters. As shown in **Fig. 1C**, genes with H3K4me3 losses in MCI and AD were enriched in synapse organization and synapse transmission; while genes with H3K4me3 gains in MCI and AD were enriched in RNA polymerase II-mediated transcription.

SYNGO analysis (**Fig. 1D**) revealed that, in MCI, the 94 synaptic genes with H3K4me3 losses were enriched in postsynaptic density, while in AD, the 119 synaptic genes with H3K4me3 losses were enriched in both postsynaptic and presynaptic sites. AD displayed more substantial H3K4me3 losses on synaptic genes than MCI.

We also generated protein-protein interaction (PPI) networks of hub genes with changed H3K4me3 in MCI and AD. For top 20 synaptic hub genes with H3K4me3 losses, MCI had voltage-gated calcium channels (VDCC) (*CACNG3* and *CACNA1A*), synaptic plasticity molecule CaMKII (*CAMK2A*), and cytoskeleton regulators Alpha-actinin-1 (*ACTN1*) and Talin-2 (*TLN2*); while AD had VDCC (*CACNG3* and *CACNG2*), synaptic organizer neurexin-1 (*NRXN1*), glutamate receptors (*GRIK2* and *GRIA2*), and synaptogamins (*SYT1* and *SYT4*) that act as Ca^2+^ sensors for neurotransmitter release (**Fig. 1E**). For the top 20 transcriptional hub genes with H3K4me3 gains, MCI had transcription factors and regulators, such as *RUNX3, GATA3*, *FOXP1*, *OTX2*, *PITX2*, *ZEB2*, *MEIS1, EZH2*, *PRDM14* and *PIK3R1,* while AD had transcription factors, including *RUNX3, GATA3, FOXA2*, *OTX2*, *SMAD3*, *NEUROD1* and *PAX2/5/8* (**Fig. 1F**).

### Module Eigengenes with Correlated H3K4me3 Binding Show Disease-Specific H3K4me3 Losses and Gains

Weighted Correlation Network Analysis (WGCNA) segregated protein-coding genes with H3K4me3 binding at their promoters into 12 module eigengenes (MEs) (**Fig. 2A**). These MEs were sorted according to their sizes, and the percentage of genes with differential H3K4me3 binding in MCI and AD (compared to controls) in each ME is shown in **Fig. 2B** (**Sup. Table 2**). Three modules were selected for more detailed analyses.

**Figure 2.**
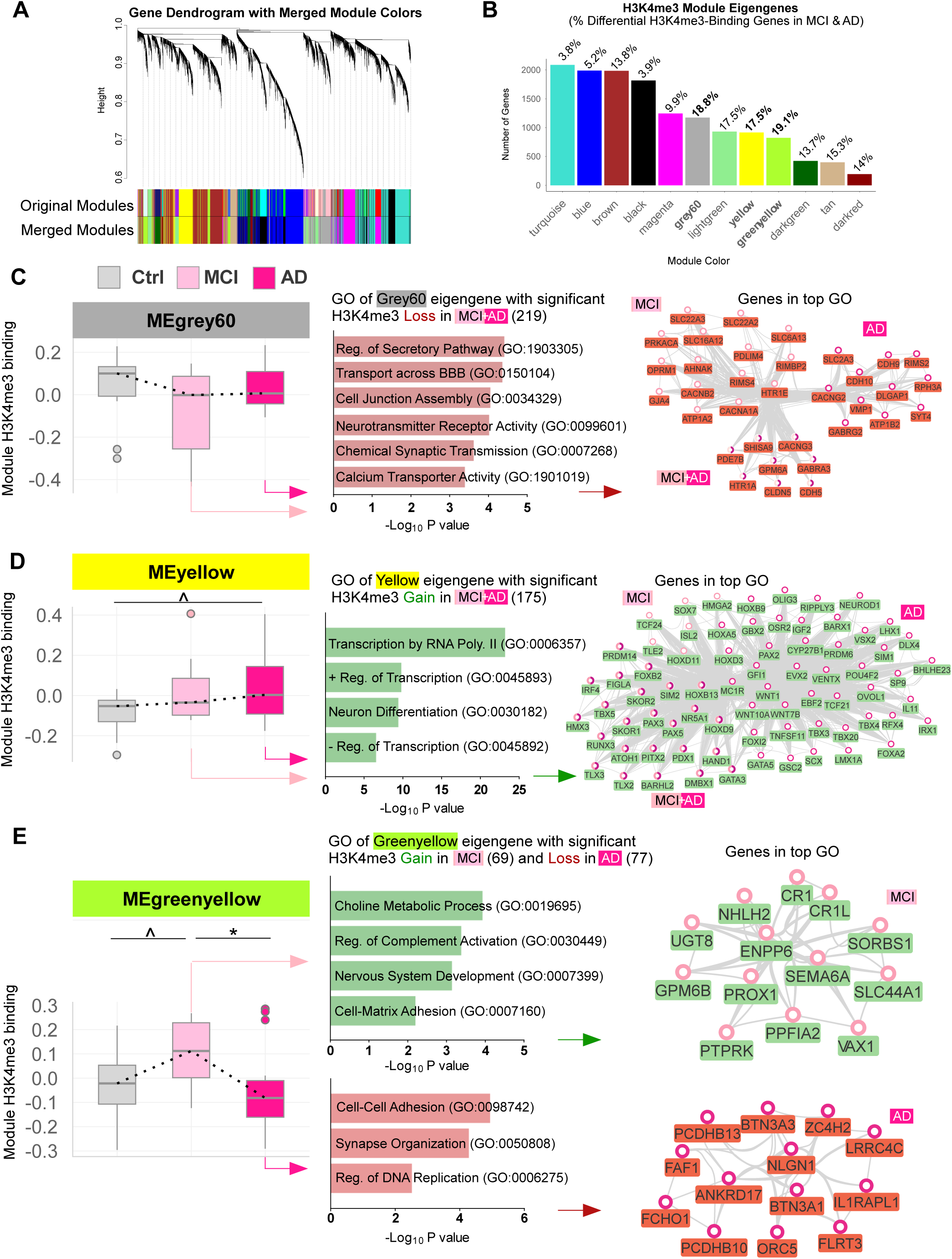
H3K4me3 Losses and Gains in Module Eigengenes with Correlated H3K4me3 Binding in MCI and AD. **A,** Dendrogram depicting protein-coding genes with H3K4me3 binding at their promoters clustered into module eigengenes (MEs) using weighted similarity scores derived from positive correlations from all 3 groups (Ctrl, MCI, AD). Modules were assigned to a random color and consolidated using a set threshold. **B,** H3K4me3 MEs sorted according to size. The percentage of genes with significant (p ≤ 0.05) H3K4me3 changes in each module is also labeled. P values were calculated using Weighted Fisher test to consolidate gene isoforms and two-sided Wilcoxon ‘Mann-Whitney’ test for individual genes. **C, F, I,** Box plots of H3K4me3 binding in MEgrey60 (C), MEyellow (F) and MEgreenyellow (I) of Ctrl, MCI, and AD (MEgrey60: p = 0.19, MEyellow: p = 0.099, MEgreenyellow: p = 0.038, one-way ANOVA with *post hoc* analysis, * p_adj._ ≤ 0.05, ^ p_adj._ ≤ 0.2). **D, G, J,** GO of MEgrey60 eigengenes with significant H3K4me3 losses in MCI and AD (D), MEyellow eigengenes with significant H3K4me3 gains in MCI and AD (G) and MEgreenyellow eigengenes with significant H3K4me3 gains in MCI and losses in AD (J). **E, H, K,** PPI of MEgrey60 (E), MEyellow (H) and MEgreenyellow (K) eigengenes in top GO with significant H3K4me3 losses or gains in MCI and AD.

MEgrey60 (1173 genes) represented the module of “Synaptic Transmission and Ion Homeostasis Regulation” and trended towards a loss of H3K4me3 binding in MCI and AD (**Fig. 2C**). The 219 MEgrey60 eigengenes with significant H3K4me3 losses in MCI and AD were enriched in regulation of secretory pathway, neurotransmitter receptor activity and chemical synapse transmission (**Fig. 2D**). PPI network of these genes in top GO (**Fig. 2E**) included VDCC (*CACNAG3/2, CACNA1A*, *CACNB2*), synaptic membrane transporters (*SLCA6A13*, *SLC16A12*, *SLC22A2*/*3)*, exocytosis regulators (*RIMS4/2*, *RIMBP2, SYT4)*, and cadherins (*CDH5/9/10)*.

MEyellow (916 genes) represented the module of “Transcriptional Regulation of Neurogenesis and Inflammatory Signaling” and trended towards an increase of H3K4me3 binding in MCI and AD (**Fig. 2F**). The 175 MEyellow eigengenes with significant H3K4me3 gains in MCI and AD were enriched in RNA polymerase II-mediated transcription, positive and negative regulation of transcription, and neuron differentiation (**Fig. 2G**). PPI network of these genes in top GO (**Fig. 2H**) included transcription factors *RUNX3*, *FOXB2*, *SOX7*, *HOXA5*, *HOXB9*, *HOXD3/9/11, PAX2/3/5* etc.

MEgreenyellow (824 genes) represented the module of “Cell Adhesion and Synaptic Organization in Neural Development” and trended towards an increase of H3K4me3 binding in MCI followed by a downward trend in AD (**Fig. 2I**). The 69 MEgreenyellow eigengenes with significant H3K4me3 gains in MCI were enriched in choline metabolic process (*UGT8* and *ENPP6)*, regulation of complement activation (*CR1*, *CR1L)*, nervous system development (*PTPRK*, *PPFIA2)* and cell-matrix adhesion (*GPM6B* and *SEMA6A)* (**Fig. 2J** and **2K**, top). The 77 MEgreenyellow eigengenes with significant H3K4me3 losses in AD were enriched in cell-cell adhesion (PCDHB*10*/*13)* and synapse organization (*NLGN1, LRRC4C*) (**Fig. 2J** and **2K**, bottom).

H3K4me3 binding in all eigengene modules (**Sup. Fig. 1**) demonstrated more complex patterns of changes of this histone mark in MCI and AD.

### H3K27me3 Occupancies are Gained on Synaptic Genes in Both MCI and AD

In addition to the permissive H3K4me3, we next examined repressive H3K27me3 ChIP-seq data in control, MCI and AD. Segregation between diseased groups and controls in the sample distribution of H3K27me3 peaks was shown in PCA plots (**Fig. 3A**). Differential analysis revealed 6650 significantly altered H3K27me3 binding sites in MCI, with much more gains (944 peaks, 844 genes) than losses (233 peaks, 217 genes) at the promoter of protein-coding genes. Similarly, in AD, 6035 changed H3K27me3 binding sites were identified with much more gains (704 peaks, 641 genes) than losses (165 peaks, 159 genes) at promoters of protein-coding genes (**Fig. 3B, Sup. Table 3**).

**Figure 3.**
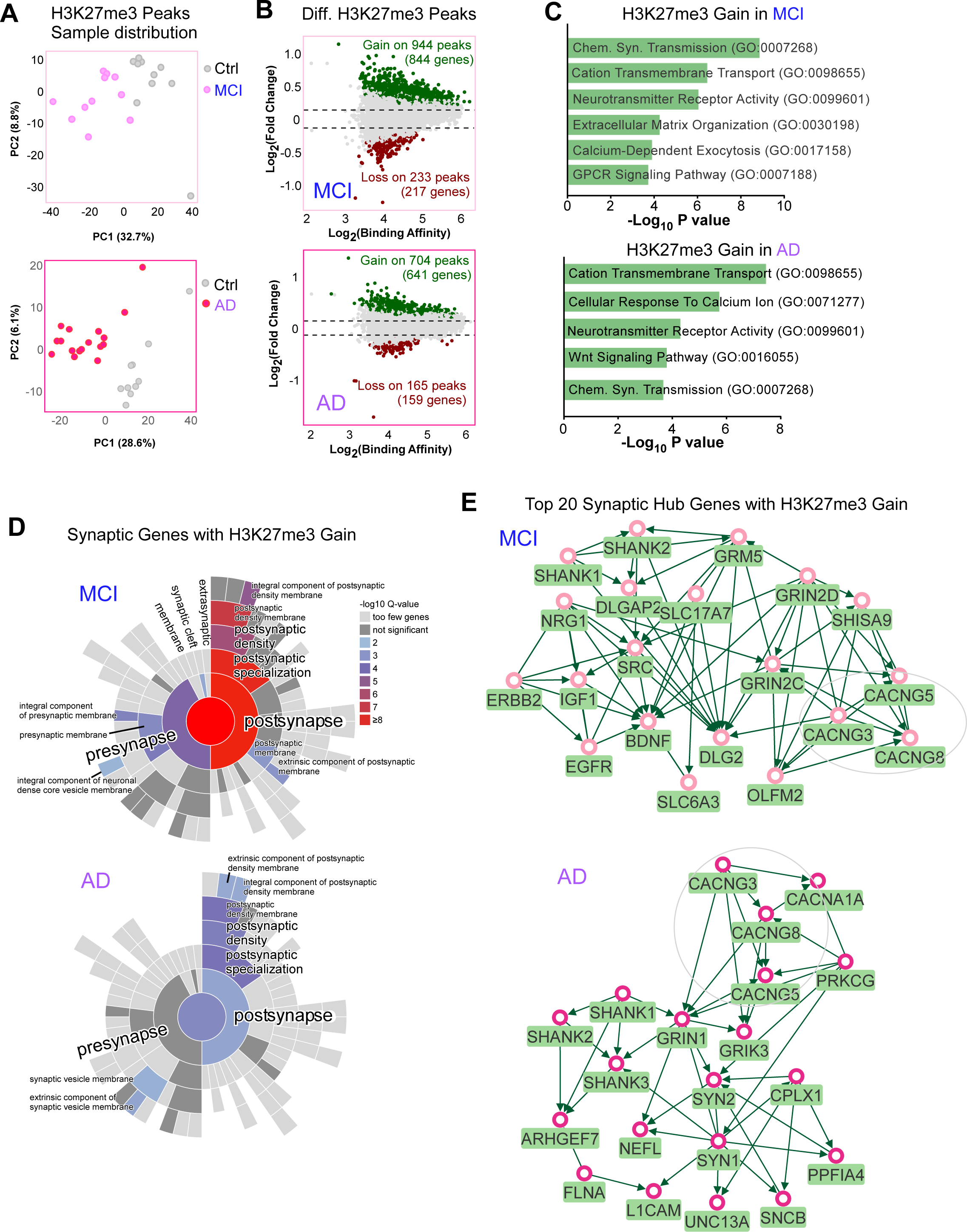
H3K27me3 Alterations in MCI and AD. **A,** PCA plots depicting H3K27me3 peaks’ sample distribution in Ctrl vs. MCI (top) or Ctrl vs. AD (bottom). **B,** MA plots depicting differential H3K27me3 binding scores in MCI (top) and AD (below), compared to Ctrl. Significant gains (green) and losses (red) are also labeled. P-values were calculated using a two-sided Wilcoxon ‘Mann-Whitney’ test, with a threshold of ≤ 0.05 **C,** GO of protein-coding genes with significant H3K27me3 gains at their promoters in MCI (top) and AD (bottom). P-values were calculated using Fisher’s Exact test. **D,** SYNGO cellular component analysis of synaptic genes with significant H3K27me3 gains in MCI (top) and AD (bottom). P-values were calculated using Fisher’s Exact test. **E,** PPI of top 20 synaptic hub genes with H3K27me3 gains in MCI (top) and AD (bottom).

In both MCI and AD, genes with significant H3K27me3 gains were enriched in chemical synapse transmission, cation transmembrane transport, and neurotransmitter receptor activity (**Fig. 3C**). SYNGO (**Fig. 3D**) revealed that the 147 synaptic genes with H3K27me3 gains in MCI were enriched in postsynaptic and presynaptic membranes, while the 87 synaptic genes with H3K27me3 gains in AD were mainly enriched in postsynaptic specializations. MCI exhibited more pronounced H3K27me3 gains on synaptic genes than AD.

As shown in PPI networks of top 20 synaptic hub genes with H3K27me3 gains (**Fig. 3E**), MCI had VDCC (*CACNG3/5/8*), glutamate synapse molecules (*GRIN2D*, *GRIN2C*, *SHANK1*/2, *GRM5, DLG2*), and growth factors (*BDNF, IGF1, EGFR*); while AD also had VDCC (*CACNG3/8, CACNA1A*), glutamate synapse molecules (*GRIN1*, *GRIK3*, *SHANK1/2/3*), and vesicle exocytosis regulators (SYN1/2, *UNC13A* and *CPLX1*).

### Module Eigengenes with Correlated H3K27me3 Binding Show Disease-Associated H3K27me3 Gains

We further performed WGCNA analyses of module eigengenes with H3K27me3 binding (**Fig. 4A**). Modules with the most significantly changed genes revealed from differential analysis were picked for further analysis (**Fig. 4B, Sup. Table 4**).

**Figure 4.**
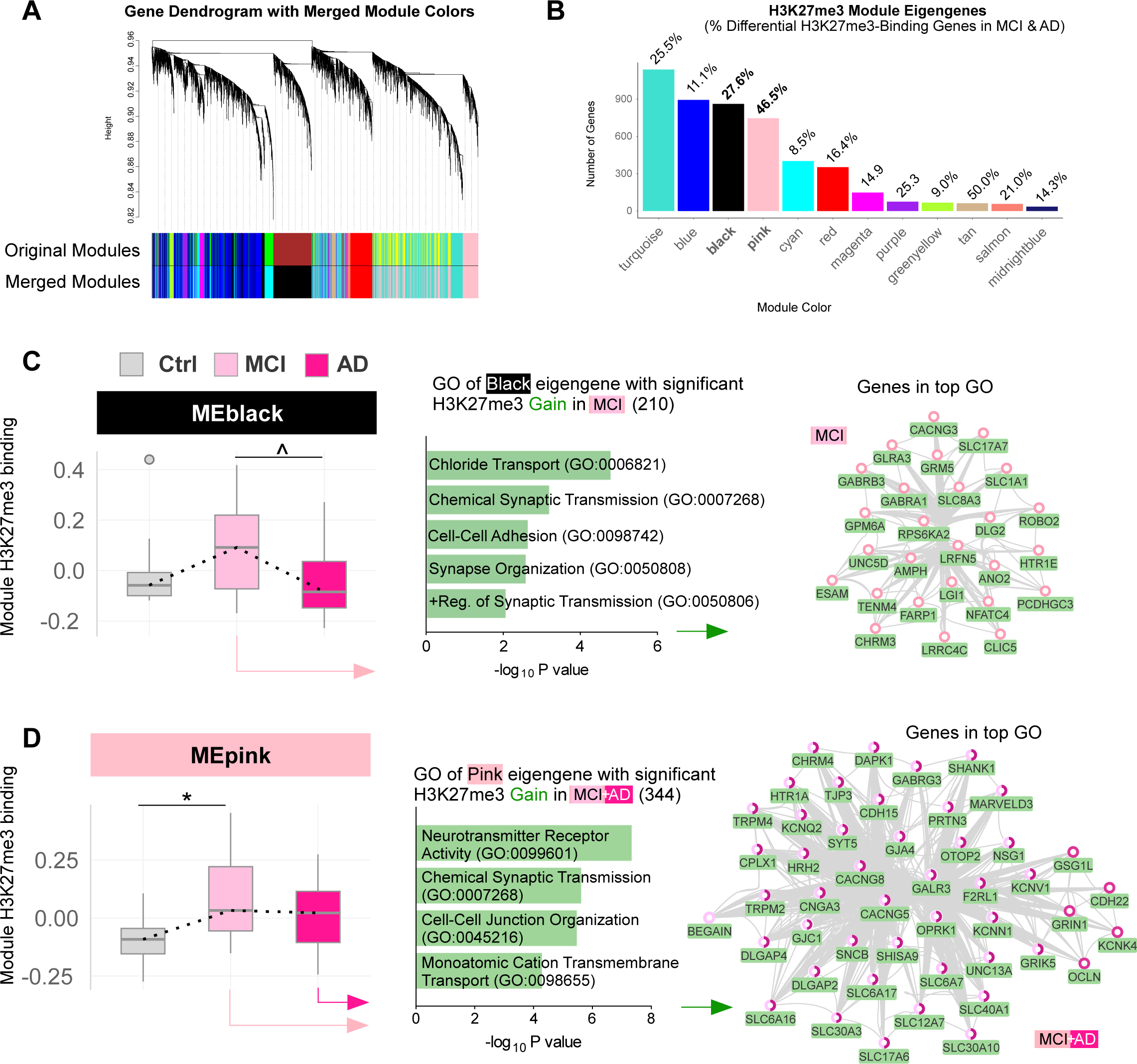
H3K27me3 Gains in Module Eigengenes with Correlated H3K27me3 Binding in MCI and AD. **A,** Dendrogram depicting protein-coding genes with H3K27me3 binding at their promoters clustered into module eigengenes (MEs) using weighted similarity scores derived from positive correlations from all 3 groups (Ctrl, MCI, AD). **B,** H3K27me3 MEs sorted according to size. The percentage of genes with significant H3K27me3 changes in each module is also labeled. **C, F,** Box plots of H3K27me3 binding in MEblack (C) and MEpink (F) of Ctrl, MCI, and AD (MEblack: p = 0.12, MEpink: p = 0.045, one-way ANOVA with post hoc analysis, * p_adj._ ≤ 0.05, ^ p_adj._ ≤ 0.2). **D, G,** GO of MEblack eigengenes with significant H3K27me3 gains in MCI (D) and MEpink eigengenes with significant H3K27me3 gains in MCI and AD (G). **E, H,** PPI of MEblack (E) and MEpink(H) eigengenes in top GO with significant H3K27me3 gains in MCI and AD.

MEblack (862 genes) represented one module of “Synaptic Signaling and Neuronal Ion Tansport” and showed a trend of increased H3K27me3 binding in MCI, but not in AD (**Fig. 4C**). The 210 MEblack eigengenes with significant H3K27me3 gains in MCI were enriched in chloride transport, chemical synaptic transmission, cell-cell adhesion, synapse organization, and synaptic transmission (**Fig. 4D**). These genes in top GO (**Fig. 4E**) included *CACNG3*, *GABRA1/3*, *DLG2*, *SLC1A1/7*, *SLC8A3*, and *PCDHGC3*.

MEpink (747 genes) represented another module of “Synaptic Signaling and Neuronal Ion Tansport” and showed a trend of increased H3K27me3 binding in both MCI and AD (**Fig. 4F**). The 344 MEpink eigengenes with significant H3K27me3 gains in MCI and AD were enriched in neurotransmitter receptor activity, chemical synapse transmission, cell-cell junction organization (**Fig. 4G**). These genes in top GO (**Fig. 4H**) included *CACNG5/8, GABRG3, SHANK1, SYT5, GRIK5,* etc.

H3K27me3 binding in all eigengene modules (**Sup. Fig. 2**) revealed additional patterns of changes of this histone mark in MCI and AD.

### Transcriptomic Changes in AD are Correlated with H3K4me3 and H3K27me3 Alterations

Given the alteration of H3K4me3 (linked to gene activation) and H3K27me3 (linked to gene repression) in AD, we examined its potential link to transcriptomic changes. Using the RNA-seq data from two large scale studies (Mayo Clinic and Mount Sinai Brain Bank), we found that downregulated genes in AD were enriched in chemical synapse transmission (**Fig. 5A**, top), similar to the GO categories of genes with significant H3K4me3 losses and H3K27me3 gains in AD (Fig. 1C and 3C). On the other hand, upregulated genes in AD were enriched in regulation of DNA-templated transcription (**Fig. 5A**, bottom), similar to the GO categories of genes with significant H3K4me3 gains (Fig. 1C).

**Figure 5.**
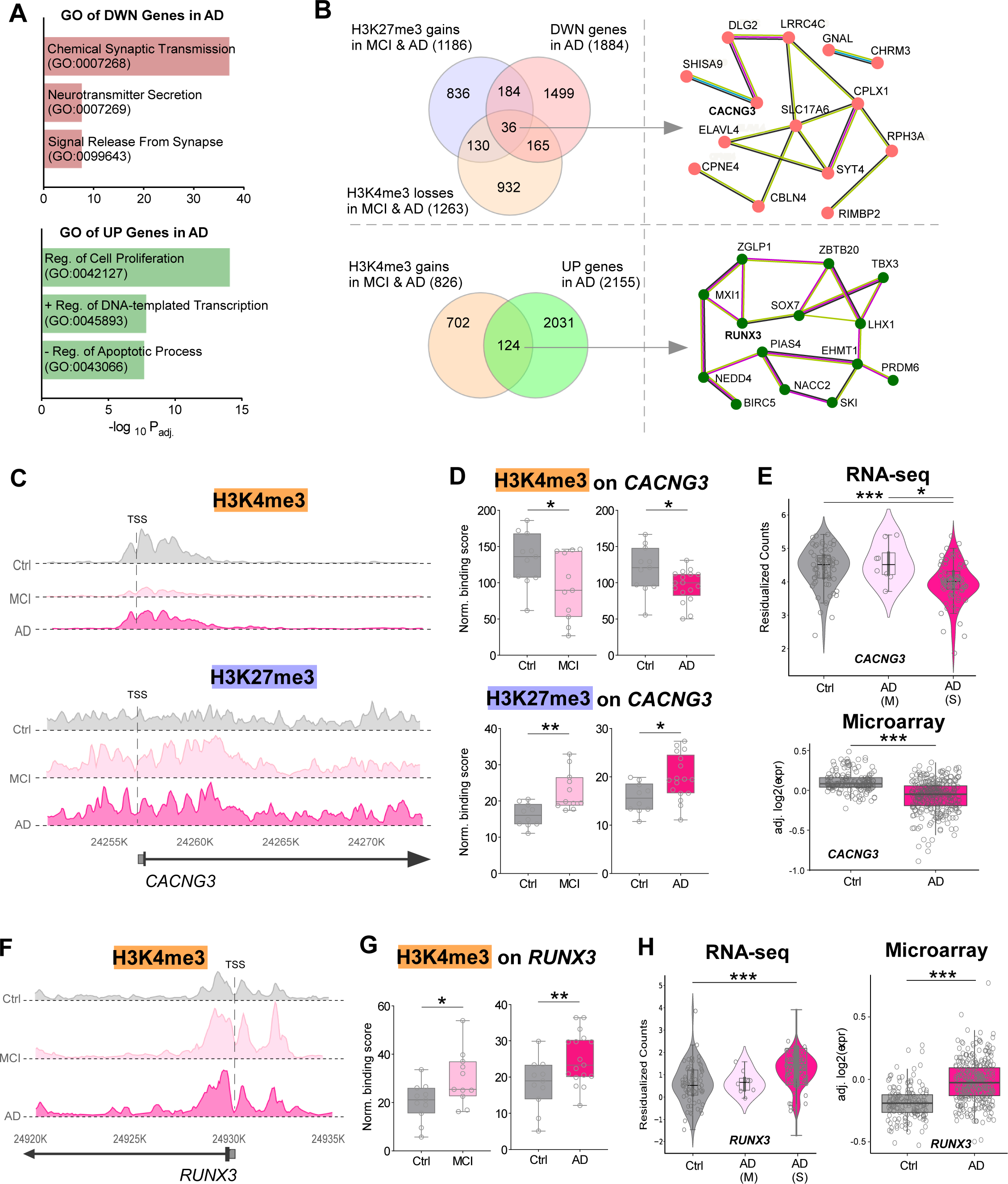
Correlation of Transcriptomic Changes in AD with H3K4me3 and H3K27me3 Alterations. **A,** GO of differentially expressed genes in AD from two large-scale RNA-seq studies (Mayo Clinic and Mount Sinai). Top: GO of downregulated genes in AD; Bottom: GO of upregulated genes in AD. P-values were calculated using Fisher’s Exact Test. **B,** Venn diagrams depicting common genes with mRNA downregulation, H3K4me3 losses and H3K27me3 gains in MCI and AD (top left), or genes with mRNA upregulation and H3K4me3 gains in MCI and AD (bottom left), as well as PPI of the common synaptic genes (top right) or transcriptional regulators (bottom right). **C, F,** ChIP-seq landscapes showing H3K4me3 and H3K27me3 peaks at *CACNG3* promoter (C) or H3K4me3 peaks at *RUNX3* promoter (F) in representative Ctrl, MCI and AD samples. **D, G,** Box plots showing H3K4me3 and H3K27me3 binding scores on *CACNG3* (H3K4me3, MCI: p = 0.038, AD: p = 0.020; H3K27me3, MCI: p = 0.002, AD: p = 0.021; Wilcoxon ‘Mann-Whitney’ test) (D) or H3K4me3 binding scores on *RUNX3* (MCI: p = 0.010, AD: p = 0.005) (G) in all Ctrl, MCI and AD samples. **E, H,** Violin and box plots of RNA-seq or microarray data showing *CACNG3* (E) or *RUNX3* (H) mRNA expression in Ctrl, moderate (M) AD or severe (S) AD groups (RNA-seq, *CACNG3*: p = 1.69e-05, *RUNX3*: p = 1.01e-04, Kruskal-Wallis test; Microarray, *CACNG3*: p = 2.2e-16, *RUNX3*: p = 2.2e-16, Wilcoxon signed-rank test).

Among the 1884 downregulated genes in AD (Mayo), 201 genes had significant H3K4me3 losses and 220 genes had significant H3K27me3 gains in MCI or AD, 36 genes showed all the three changes (transcriptional down, H3K4me3 loss, H3K27me3 gain) **(Fig. 5B, top left)**, 18 of which were synaptic genes including *CACNG3* (VDCC) (**Fig. 5B, top right, Sup. Table 5**).

On the other hand, among the 2155 upregulated genes in AD (Mayo), 124 genes had significant H3K4me3 gains in MCI or AD **(Fig. 5B, bottom left)**. These genes were highly enriched in DNA-templated transcription regulation, including *RUNX3* (transcription factor) (**Fig. 5B, bottom right, Sup. Table 5**).

We selected *CACNG3* and *RUNX3* for further analyses. As shown in the representative ChIP-seq landscape plots (**Fig. 5C**), in both MCI and AD, H3K4me3 peaks at *CACNG3* promoter were markedly reduced, and H3K27me3 peaks at *CACNG3* promoter were pronouncedly increased. Statistical analyses of all samples revealed the significant loss of permissive H3K4me3 binding and gain of repressive H3K27me3 binding on *CACNG3* in MCI and AD (**Fig. 5D**), correlated with the significant reduction of *CACNG3* mRNA expression in AD by RNA-seq and microarray studies (**Fig. 5E**). Similarly, ChIP-seq also showed the significantly increased H3K4me3 binding at *RUNX3* promoter (**Fig. 5F, G**), which was correlated with the significantly increased *RUNX3* mRNA expression in AD from RNA-seq and microarray data (**Fig. 5H**).

## Discussion

Our study demonstrates significant alterations in histone methylation landscapes in the prefrontal cortex during the progression from MCI to AD, highlighting critical epigenetic events linked to transcriptional dysregulation. Specifically, we identified robust reductions of permissive H3K4me3 and concomitant increases of repressive H3K27me3 marks at promoters of synaptic genes, accompanied by heightened H3K4me3 occupancies at transcriptional regulators. These findings suggest a coordinated epigenomic reorganization that underlies synaptic dysfunction, a hallmark of AD pathology (DeKosky & Scheff 1990, Selkoe 2002, Terry et al 1991). We propose a model in which progressive histone modification alterations contribute to transcriptional dysregulation, synaptic loss, and cognitive impairment (**Fig. 6**).

**Figure 6.**
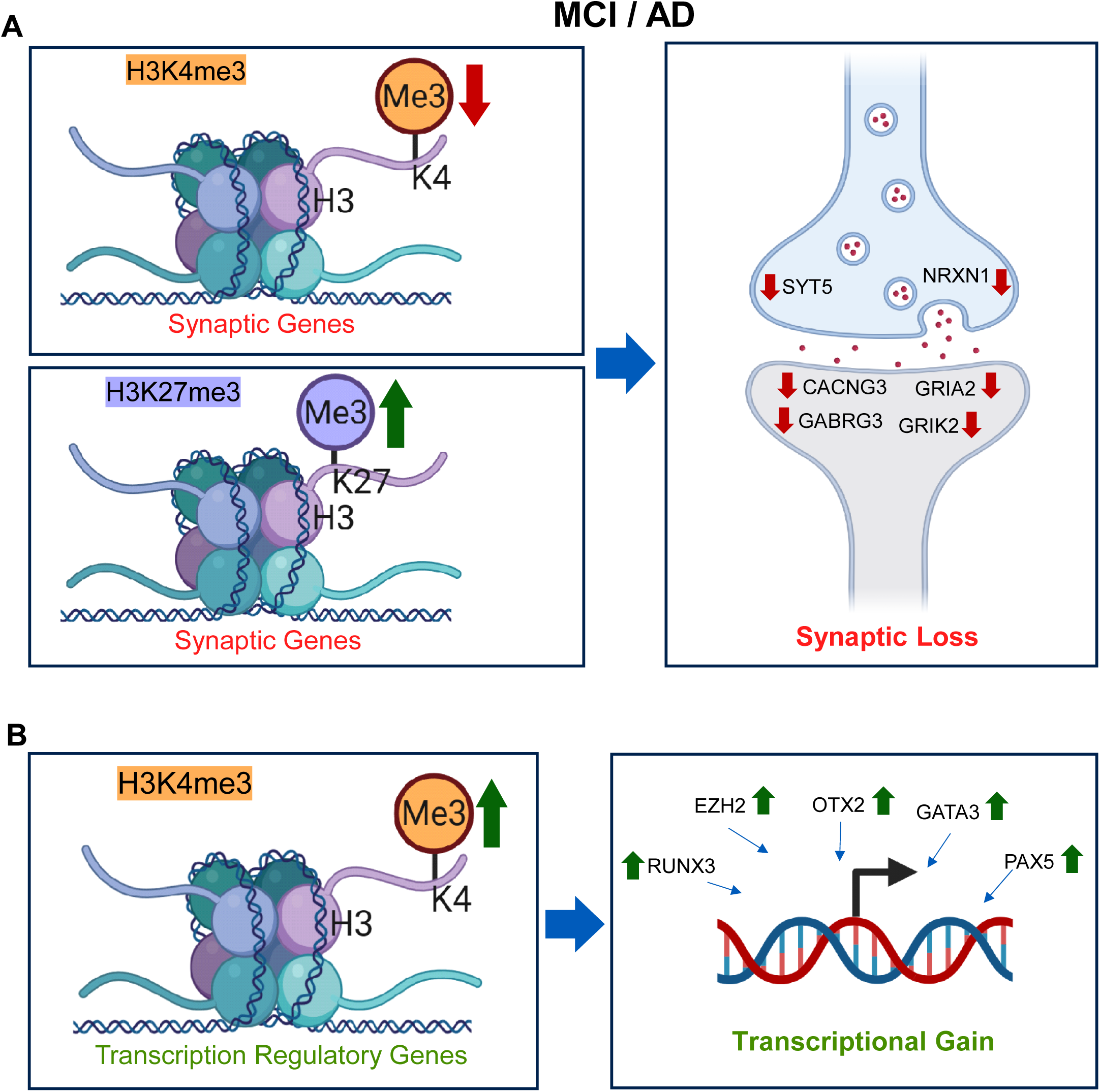
A Schematic Model of Epigenomic Changes and Consequences in MCI and AD. **A,** The loss of permissive H3K4me3 and gain of repressive H3K27me3 on synaptic genes in MCI and AD lead to the reduced expression of synaptic genes, causing the diminished synaptic function. **B,** The gain of permissive H3K4me3 on transcription regulatory genes in MCI and AD leads to excessive transcriptional activity, causing the aberrant gene regulation.

In both MCI and AD, synaptic genes, such as *CACNG3, SYT5, NRXN1, GRIA2, GABRG3,* and *GRIK2,* exhibit diminished H3K4me3 and increased H3K27me3, which synergistically reduces their transcription, leading to the impaired neuronal plasticity, neurotransmitter signaling, and cognitive function (Gräff & Tsai 2013, Peixoto & Abel 2013). As shown in one example, *CACNG3* (a VDCC subunit), its pronounced H3K4me3 depletion and robust H3K27me3 enrichment in MCI and AD correlate strongly with transcriptional downregulation in AD. Dysfunction of VDCCs, particularly CACNG subunits, disrupts calcium homeostasis critical for synaptic plasticity, memory formation, and neuronal survival (Seoane et al 2009, Simms & Zamponi 2014). Thus, epigenetic suppression of *CACNG3* could directly contribute to synaptic dysfunction in AD pathogenesis.

Our findings align well with a genome-wide characterizations of histone methylation in AD brains, which showed a loss of H3K4me3 at genes critical for synaptic function (Persico et al 2022). Interestingly, familial AD models also show altered histone methylation, predominantly H3K4me3 losses at neuronal identity genes and H3K27me3 gains at synaptic loci (Caldwell et al 2020). These changes closely mirror our findings in sporadic AD, suggesting a unified epigenetic vulnerability potentially driving cognitive decline irrespective of genetic etiology.

In parallel, we observed significant increases of H3K4me3 occupancies at promoters of transcriptional regulators, such as *RUNX3, EZH2, OTX2, GATA3,* and *PAX5*, leading to elevated transcriptional activity. These transcription factors orchestrate cellular responses, including DNA repair, neuronal differentiation, and inflammation (Levanon et al 2002, Tsarovina et al 2010, Yagi et al 2010). As shown in one example, *RUNX3,* its increased H3K4me3 occupancy in MCI and AD correlate strongly with transcriptional upregulation in AD. RUNX family transcription factors have been recognized as neuroprotective modulators that enhance DNA repair and survival pathways in neurodegenerative contexts (Dutta & Osato 2023, Tay et al 2018). Increased expression of such transcriptional regulators may thus represent a compensatory response to neurodegenerative stress, potentially aimed at mitigating cellular damage and neuronal loss characteristic of AD.

The observed greater loss of permissive H3K4me3 at synaptic genes in AD compared to MCI, and conversely, the more pronounced gain of repressive H3K27me3 in MCI compared to AD, likely reflect distinct stages of epigenetic and neuronal adaptation during disease progression. Early in disease (MCI), the enhanced deposition of H3K27me3 at synaptic gene promoters may represent an actively regulated neuronal stress response, serving as a potentially reversible epigenetic mechanism aimed at transiently reducing synaptic load and metabolic demands. As pathology advances into AD, persistent neuronal insults, such as chronic inflammation, amyloid accumulation, and tau pathology, may overwhelm these protective mechanisms, resulting in permanent chromatin remodeling marked by pronounced loss of H3K4me3. Consequently, the severe reduction of permissive H3K4me3 on synaptic genes in AD likely indicates irreversible transcriptional dysfunction, reflecting advanced neuronal and synaptic deterioration characteristic of late-stage neurodegeneration.

Our observation that histone methylation alterations occur in a disease stage-dependent manner highlights the complexity of chromatin remodeling in AD progression. WGCNA analyses emphasize the nuanced epigenetic dynamics: some modules showed H3K4me3 losses or gains throughout disease progression (e.g., MEgrey60 and MEyellow), whereas some modules had H3K4me3 gains in MCI but losses in AD (e.g., MEgreenyellow). Such dynamic chromatin state changes likely reflect evolving responses to neuronal stress and shifting transcriptional programs as degenerative signals proceed (Klein et al 2019, Nativio et al 2020).

Given the mechanistic significance of histone modifications, targeting dysregulated chromatin states could represent a novel therapeutic avenue. In agreement with this, pharmacological manipulation of epigenetic enzymes to normalize histone methylation (H3K4me3 and H3K9me2) levels ameliorated cognitive and synaptic deficits in mouse models of AD (Cao et al 2020, Wang et al 2021, Williams et al 2023, Zheng et al 2019).

In summary, our comprehensive profiling of histone methylation dynamics highlights critical stage-dependent epigenetic reorganization events influencing synaptic dysfunction and transcriptional responses in MCI and AD. Our findings, synergizing with recent literature, reinforce the importance of histone modification changes in neurodegenerative progression and open novel therapeutic strategies targeting chromatin dysregulation at early stages of disease.

## Methods

### Human Samples

ChIP-seq data of controls (H3K4me3: n = 13; H3K27me3: n = 12), MCI (H3K4me3 and H3K27me3: n = 11), and AD (H3K4me3 and H3K27me3: n = 17) samples (all females) were acquired from the Rush Alzheimer’s Disease Study uploaded on the Encode Project Consortium https://www.encodeproject.org (Dunham et al 2012, Luo et al 2020). Outliers (2 controls for H3K4me3, 2 controls for H3K27me3) determined by principal component analysis were removed before further analyses. Demographic information of the donors is included in **Sub Table 6**.

### Bioinformatics Analysis

Data matrices were processed in python before proceeding with analyses. Both MCI and AD groups were normalized separately for each histone modification target along with matching controls using Post Trimmed Mean of M-values (TMM) normalization implemented using “edgeR” (Robinson et al 2010) in the “DiffBind” R package (v. 3.16) (Ross-Innes et al 2012).

Weighted Correlation Network Analysis (WGCNA) (Langfelder & Horvath 2008) was used to segregate protein-coding genes with H3K4me3 or H3K27me3 binding at promoters in all 3 groups (Ctrl, MCI and AD) into 12 module eigengenes (MEs) according to a positive correlation similarity matrix and assigned a randomized color using a softpower of 9 for H3K4me3 and 5 for H3K27me3 “WGCNA” R package (v. 1.73). Correlation networks were generated via importing edge connections into Cytoscape (Shannon et al 2003) (v. 3.10.x). All Gene Ontology analyses were performed using “EnrichR” (Chen et al 2013). Protein-protein interaction networks were generated using string database (Szklarczyk et al 2021) (String-db.org), exported into “Cytoscape”. All plots were generated mainly in R using the “ggplot2” package (Wickham 2016), along with other supporting R packages.

## Acknowledgements

We thank University at Buffalo’s Center for Computational Research. This work is supported by grants from NIH (R01-AG064656 and R01-AG067597) and VA (I01-BX006357) to Z.Y.

## Data Availability

ChIP-seq data was acquired from ENCODE. Experiment accession numbers are included in **Sup. Table 6**. Total-RNA -seq data for comparisons were acquired from Mount Sinai Brain Bank (MSBB) (Wang et al 2018) and Mayo Clinic (Allen et al 2016) and the analysis from the Harmonization Study (Synapse IDs: syn3159438, syn5550404, syn9702085). Microarray data was acquired from Icahn School of Medicine at Mount Sinai’s microarray study (Zhang et al 2013) (GEO accession number: GSE44772).

**Sup. Figure 1** Box plots of H3K4me3 binding in all modules of Ctrl, MCI, and AD.

**Sup. Figure 2** Box plots of H3K27me3 binding in all modules of Ctrl, MCI, and AD.

**Sup. Table 1** Protein-coding genes with significant H3K4me3 gains and losses at their promoters in MCI and AD.

**Sup. Table 2** Module eigengenes composition with significant H3K4me3 changes in MCI or AD.

**Sup. Table 3** Protein-coding genes with significant H3K27me3 gains at their promoters in MCI and AD.

**Sup. Table 4** Module eigengenes composition with significant H3K27me3 changes in MCI or AD.

**Sup. Table 5** Genes with changes in H3K4me3, H3K27me3 and mRNA expression in MCI and AD.

**Sup. Table 6** Demographic information and accession numbers of samples.

