## Supplementary material for "Epigenomic Aberrations of Histone Methylation in Prefrontal Cortex of Humans with Mild Cognitive Impairment and Alzheimer’s Disease": Sup. Figures

Sup. Fig. 1

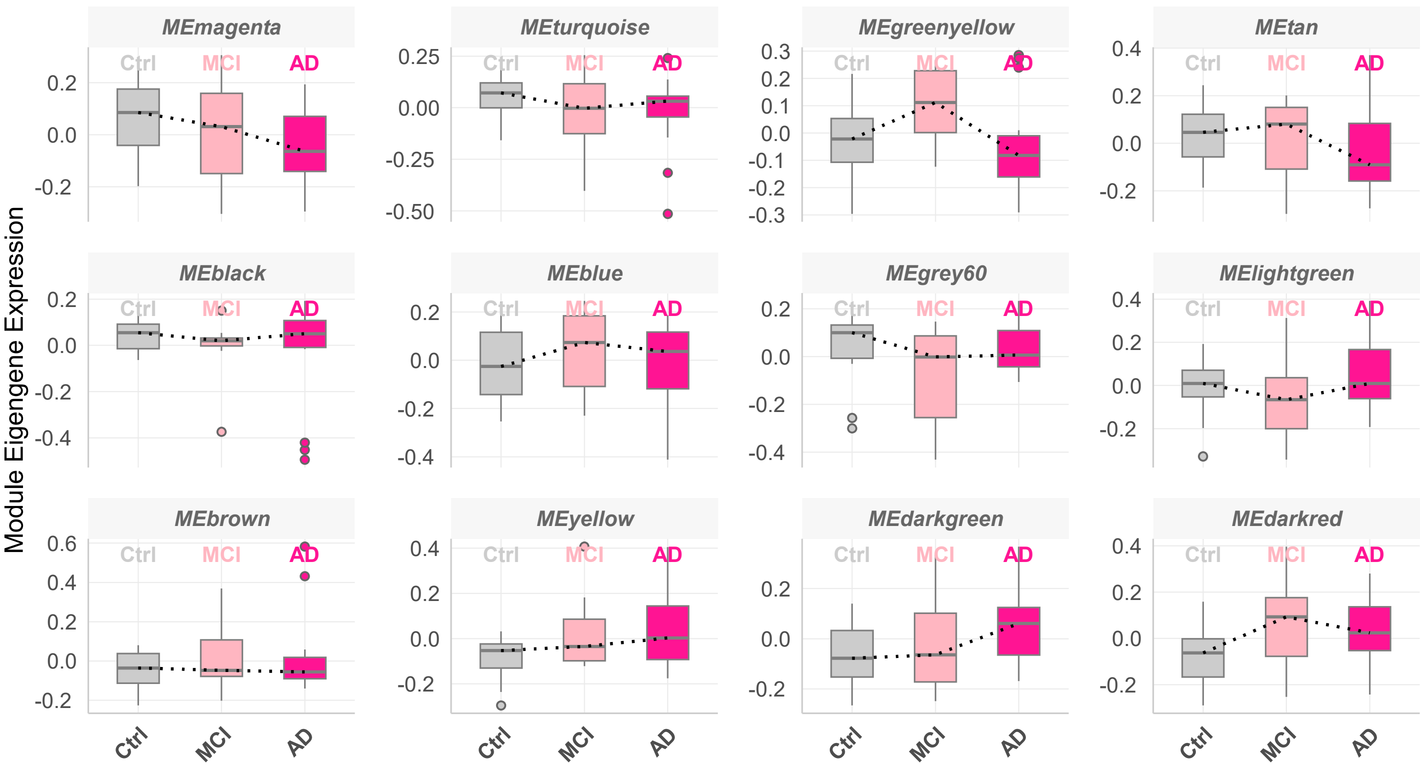


H3K4me3 Module Eigengene Expression with Disease Progression

**Sup. Fig. 2**

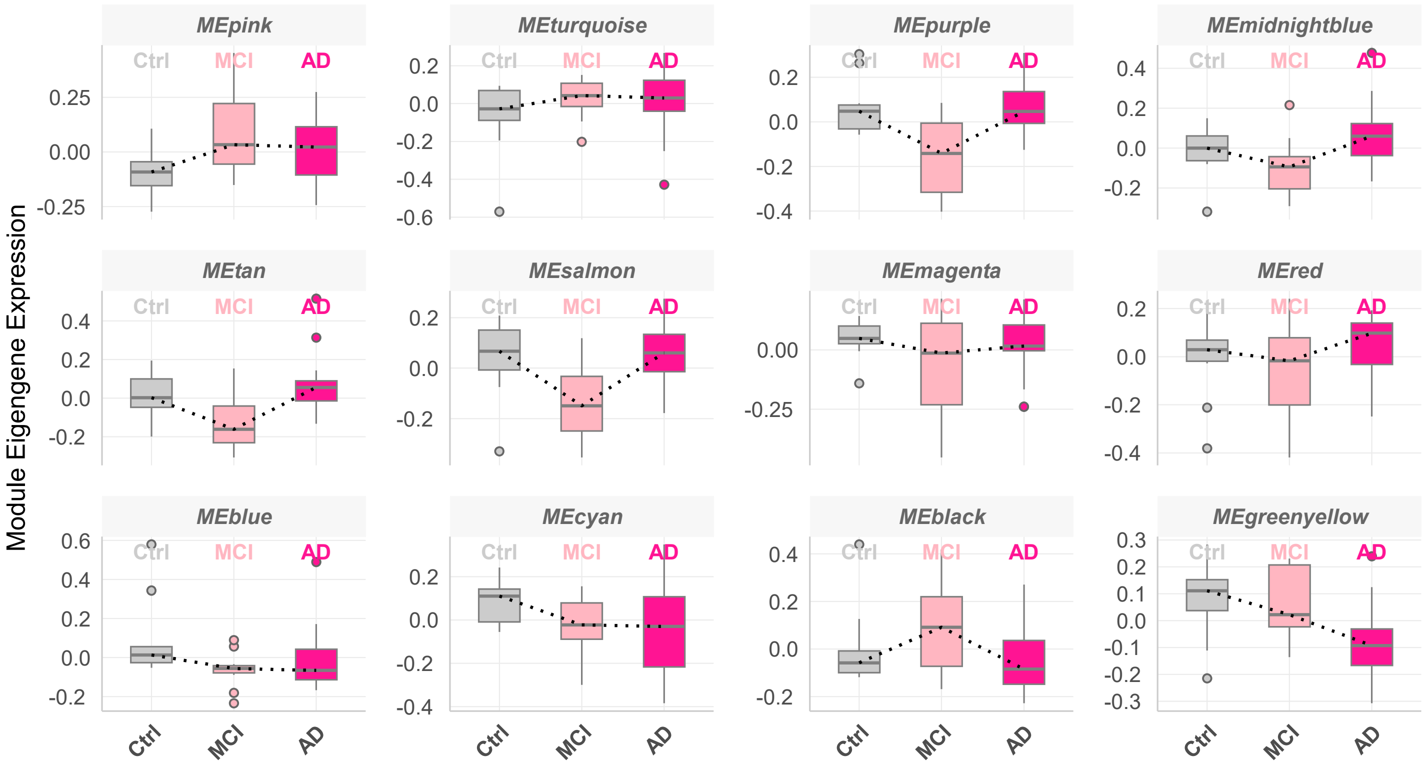


**H3K27me3 Module Eigengene Expression with Disease Progression**
